# Antibody landscape against SARS-CoV-2 proteome revealed significant differences between non-structural/ accessory proteins and structural proteins

**DOI:** 10.1101/2020.12.08.20246314

**Authors:** Yang Li, Zhaowei Xu, Qing Lei, Dan-yun Lai, Hongyan Hou, He-wei Jiang, Yun-xiao Zheng, Xue-ning Wang, Jiaoxiang Wu, Ming-liang Ma, Bo Zhang, Hong Chen, Caizheng Yu, Jun-biao Xue, Hai-nan Zhang, Huan Qi, Shu-juan Guo, Yandi Zhang, Xiaosong Lin, Zongjie Yao, Huiming Sheng, Ziyong Sun, Feng Wang, Xionglin Fan, Sheng-ce Tao

## Abstract

The immunogenicity of SARS-CoV-2 proteome is largely unknown, especially for non-structural proteins and accessory proteins. Here we collected 2,360 COVID-19 sera and 601 control sera. We analyzed these sera on a protein microarray with 20 proteins of SARS-CoV-2, built an antibody response landscape for IgG and IgM. We found that non-structural proteins and accessory proteins NSP1, NSP7, NSP8, RdRp, ORF3b and ORF9b elicit prevalent IgG responses. The IgG patterns and dynamic of non-structural/ accessory proteins are different from that of S and N protein. The IgG responses against these 6 proteins are associated with disease severity and clinical outcome and declined sharply about 20 days after symptom onset. In non-survivors, sharp decrease of IgG antibodies against S1 and N protein before death was observed. The global antibody responses to non-structural/ accessory proteins revealed here may facilitate deeper understanding of SARS-CoV-2 immunology.

**Highlights:**

- An antibody response landscape against SARS-CoV-2 proteome was constructed
- Non-structural/accessory proteins elicit prevalent antibody responses but likely through a different mechanism to that of structural proteins
- IgG antibodies against non-structural/accessory proteins are more associated with disease severity and clinical outcome
- For non-survivors, the levels of IgG antibodies against S1 and N decline significantly before death

## Introduction

COVID-19, caused by SARS-CoV-2 ^1,2^, has become one of the most threatening crisis to global public health. By November 4, 2020, 47,328,401 cases were diagnosed and 1,212,070 lives were claimed (https://coronavirus.jhu.edu/map.html)^3^. SARS-CoV-2 belongs to the betacoronavirus genus and its genome encodes four major structural proteins, *i. e*., spike (S), envelope (E), membrane (M), and nucleocapsid (N), and 15 non-structural proteins (Nsp1-10 and Nsp12-16) and 9 accessory proteins^4^. Among them, the S protein, consists of N-terminal S1 fragment and C-terminal S2 fragment, plays an essential role in viral attachment, fusion, and entry into the target cells that express the viral receptor, *i. e*., angiotensin-converting enzyme 2 (ACE2)^5–9^. While the function, including immunogenicity of most of the non-structural proteins and accessory proteins are still elusive.

One of the major features of COVID-19 patients is the extreme variability of clinical severity from asymptom to death^10^. However, the factors that cause this variability are still largely unknown. Humoral immune responses elicited by SARS-CoV-2 play essential roles, especially in diagnosis, neutralizing antibody production and vaccine development^11–13^. Among all the SARS-CoV-2 proteins, S protein and N protein exhibit high immunogenicity. Antibodies against S protein and N protein are elicited in most patients, and with higher titers in severe patients, demonstrating the association between severity and humoral immune responses ^12,14^. It was reported that the antibodies against peptides derived from non-structural and accessory proteins were also detectable in patients ^10,15,16^. However, the prevalence, clinical relevance and the dynamic of non-structural proteins and accessory proteins in patients are still largely unknown.

Recently, we constructed a SARS-CoV-2 proteome microarray, containing S protein, N protein and most of the NSPs and accessory proteins^14^. The microarray is a powerful tool to systematically study the humoral immune response, especially the IgG and IgM responses against the SARS-CoV-2 proteome. Based on this platform, we have successfully characterized the humoral immune in convalescent patients^14^, asymptomatic patients^17^.

Here, we adopted an updated SARS-CoV-2 proteome microarray that contains 20 proteins, profiled 2,360 sera from 783 COVID-19 patients and 601 control sera. We identified that NSP1, NSP7, NSP8, RdRp, ORF3b and ORF9b which can strongly elicit antibodies in COVID-19 patients. Further analysis revealed that the patterns of humoral immunity of these non-structural/accessory proteins were distinct from that of S and N protein. The global antibody responses to non-structural proteins and accessory proteins revealed in this study will facilitate the comprehensive understanding of SARS-CoV-2 humoral immunity, and may provide potential biomarkers for precise monitoring of COVID-19 progression.

## Results

### COVID-19 severity and clinical outcome are associated with a set of clinical parameters

To systematically analyze the clinical characteristics of SARS-CoV-2 infection, we first analyzed the correlations between severity and each of the available laboratory parameters of 783 COVID-19 patients monitored when admission (**Table S1**). According to the severity and clinical outcome, the patients were divided into three groups, *i. e*., Non-severe patients, all of whom were recovered, severe but survived patients and non-survivors. Statistical comparison among these three groups enables us to investigate the features either related to the severity for survivors or to the outcome under similar severity (**Table 1**). Since for some patients some laboratory examinations were missing, for each clinical parameter only the effective patient numbers were given. Expectedly, gender, age, comorbidities of hypertension and diabetes are associated factors of severity, however, only age are significantly associated with clinical outcome for severe patients. In addition, in consistent with many previous studies^18–20^, we identified a set of clinical and laboratory parameters which are highly related to severity or outcome, such as lymphopenia, increased CRP (C reaction protein) and factors associated with blood coagulation, cardiac injury, liver injury and kidney injury. Most of these factors are associated both with severity and outcome, while some are likely associated either with severity or outcome. For instance, thrombocytopenia and some kidney injury related factors are more common in non-survivors as compared to the other two groups, while most liver injury related factors are only associated with severity but not outcome.

**Table 1.**
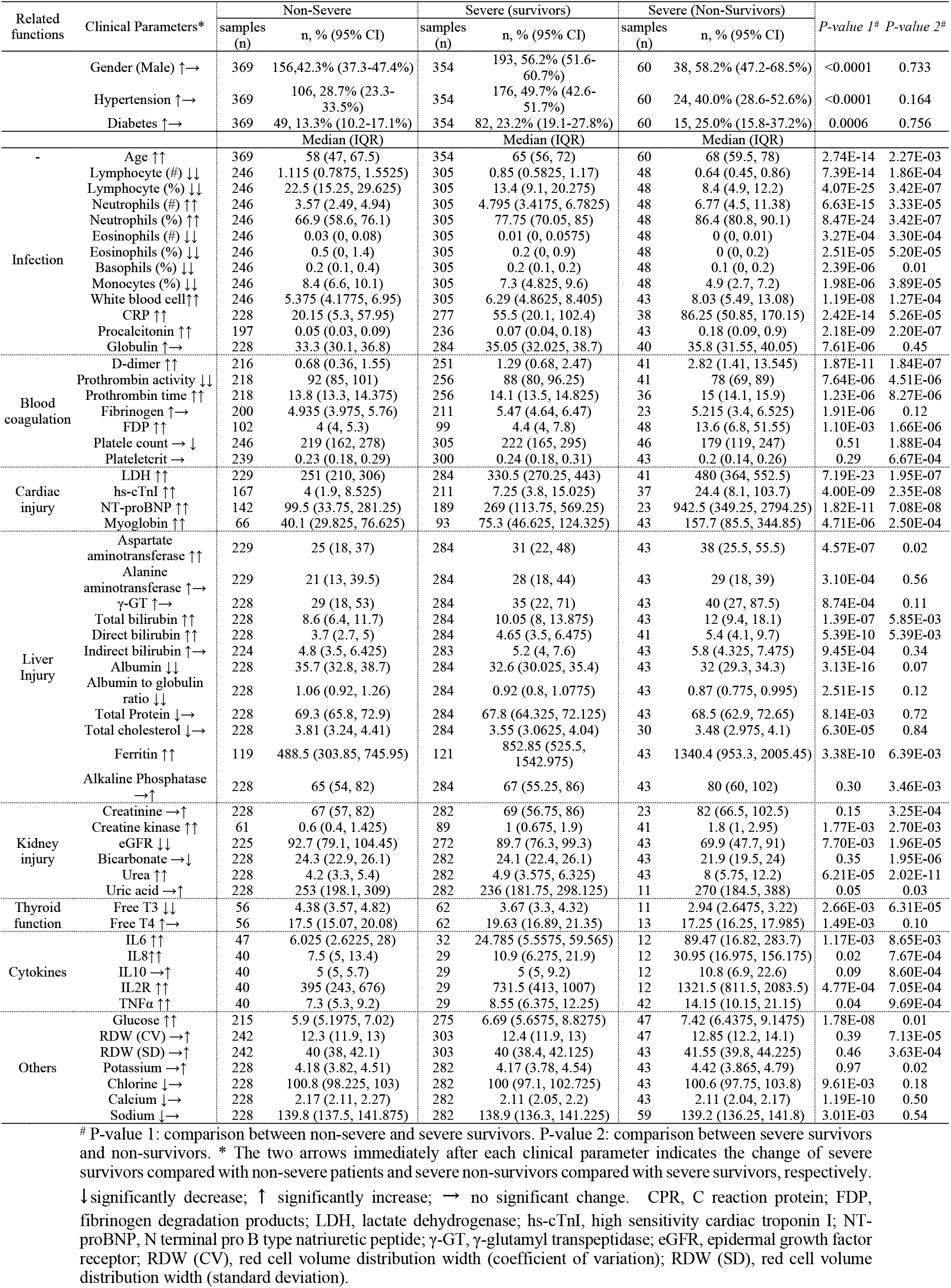
Clinical Parameters related to severity of COVID-19 patients.

### Several non-structural and accessory proteins elicit highly prevalent antibody responses

Previously, we have constructed a SARS-CoV-2 proteome microarray (**Figure S1a, b**) and screened a small cohort of convalescent patients ^14^. Here, we aimed to systematically analyze the immune responses and its dynamic change against SARS-CoV-2 proteins with a much larger cohort of samples. In total, we collected 2,360 sera from 783 laboratory confirmed COVID-19 patients as well as 601 control sera (**Table S1**). All of these sera were analyzed on the SARS-CoV-2 protein microarray. To acquire high-quality data for the microarray experiments, we prepared a positive control by mixing 50 randomly selected COVID-19 sera. This control was then probed on each microarray to assess and normalize the data. It turned out that high reproducibility was achieved in our assa (**Figure S1c, d**). To simplify the analysis and assure the comparability among different SARS-CoV-2 proteins, we defined “initial serum” as the first serum collected 14 days after symptom onset for each patient. The results of the initial sera were used to construct the antibody response landscape (**Figure 1**). Immune response frequency was calculated for each protein with the cutoff value set by mean + 2 x SD of the control group. Except for S1 and N, which are known of highly antigenic, we found that several non-structural and accessory proteins elicited prevalent antibody responses, especially for IgG, including NSP1, NSP7, NSP8, RdRp, ORF3b and ORF9b, for which the positive rates are 38%, 48.4%, 27.9%, 30.3%, 52.1% and 28%, respectively. Although the IgM responses were high in some cases, the overall responses are much lower than that of IgG. We then decided to focus on IgG for in-depth analysis.

**Figure 1.**
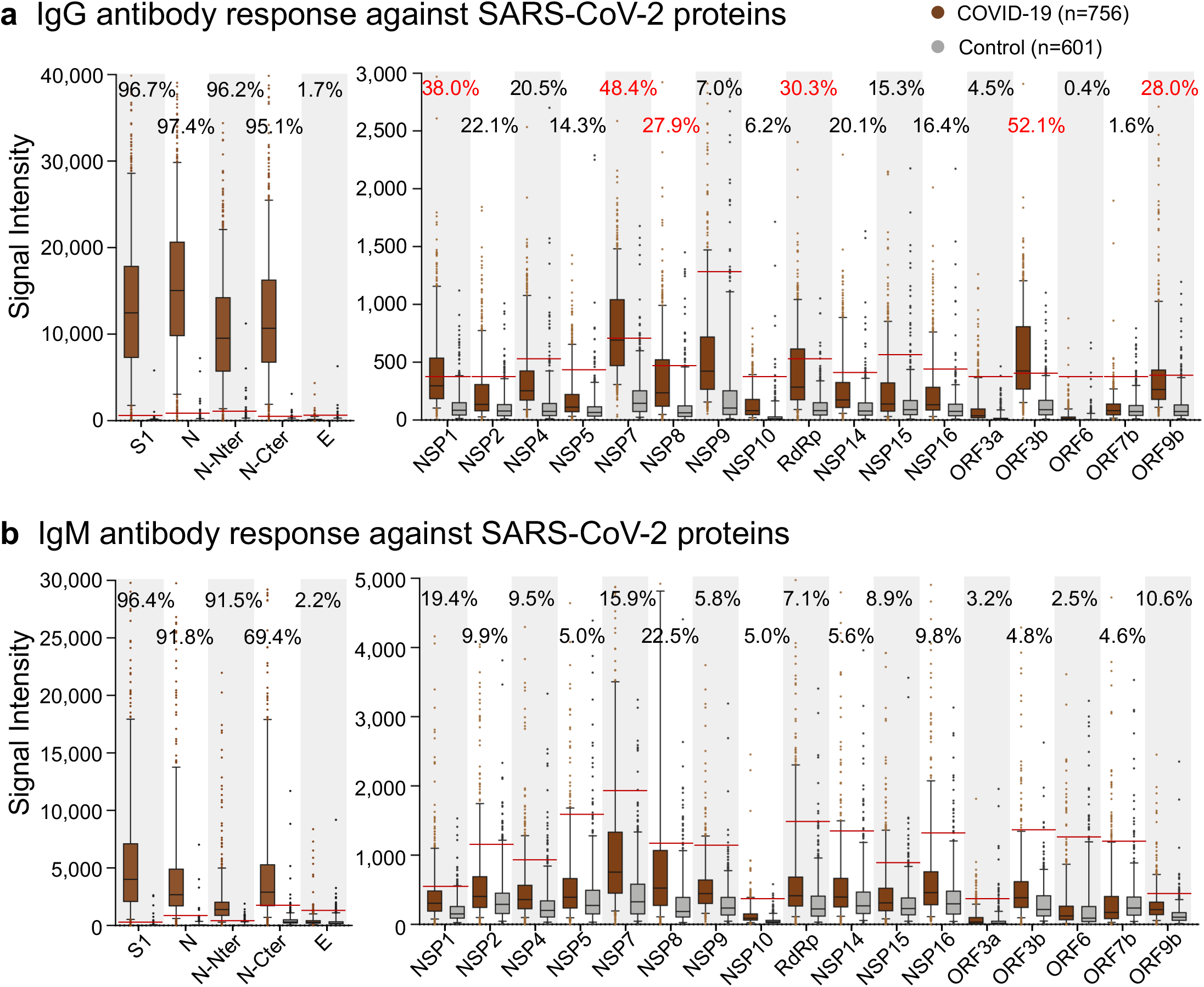
Antibody response landscape against SARS-CoV-2 proteins. IgG (**a**) and IgM (**b**) responses against each SARS-CoV-2 protein was depicted as boxplot according to the signal intensity of each sample on the proteome microarray. The data were presented as median with quintiles and the hinges (n = 756). Cutoff values (the red line) for each protein were set as mean + 2 x SD of the control group (n = 601), the positive rates of the patient group were labeled for each protein, positive rates > 25% are labeled as red.

### The IgG pattern of Non-structural and accessory proteins is distinct from that of S1 and N protein

We next asked whether the IgG responses to these proteins are associated with each other. We chose NSP7 as an example. The samples were divided into two groups depending on positive or negative of NSP7 IgG. Positive rates of the rest proteins were calculated for the two groups. Unexpectedly, for all the non-structural and accessory proteins, except for ORF3a, ORF6 and ORF7a which barely elicit antibodies, the positive rates in NSP7-IgG positive group was significantly higher than that in NSP7-IgG negative group, demonstrating high correlations (**Figure 2a**). Interestingly, there is no obvious difference for the IgG responses of S1 and N. To further confirm our observation, we reversely compared the positive rates of IgG to non-structural/accessory proteins between the groups of S1-IgG positive and negative (**Figure 2b**). The positive rates of IgG response of N protein are significantly different between these two groups, while no obvious difference was shown for the non-structural/accessory proteins. These observations demonstrate that the structural proteins elicit antibodies with distinct pattern to that of the non-structural and accessory proteins, suggesting that the underlying mechanisms by which the antibodies are triggered are different for these two groups of proteins. To further study the correlations of IgG signal intensity among the proteins, Pearson correlation coefficients between any two of these proteins were calculated and then clustered. The proteins with less than 10% response frequency were not included duo to statistical limit. (**Figure 2c**).

**Figure 2.**
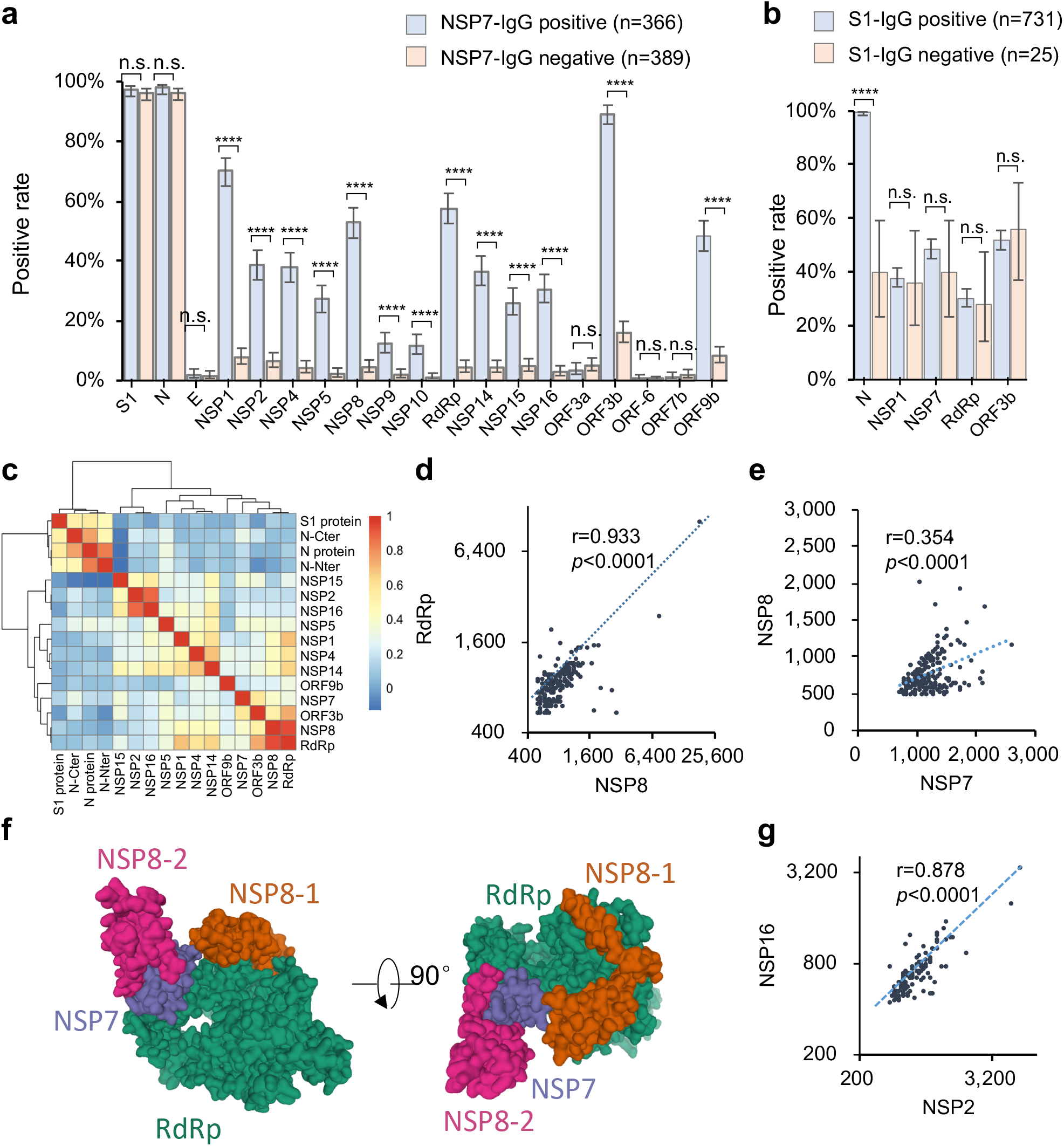
Antibodies against structural proteins and other proteins are in different patterns. **a**. Antibody positive rates for the SARS-CoV-2 proteins in two patient groups, the patient groups were divided according to NSP7 IgG signal, either positive or negative. **b**. Antibody positive rates for selected proteins in two patient groups, the patient groups were divided according to S1 IgG signal, either positive or negative. **c**. The Pearson correlation coefficients of the IgG responses among the proteins were calculated and clustered. **d-e**. Correlations of the IgG responses against RdRp and NSP8 (**d**), NSP8 and NSP7 (**e**). **f**. The location and accessibility of NSP7, NSP8 and RdRp in the SARS-CoV-2 RNA polymerase complex (PDB: 7BV1). **g**. Correlations of the IgG responses against NSP2 and NSP16. For **a-b**, error bar was given as the 95% confidential interval. *P*-value was calculated by two-sided χ2 test. *, P < 0.05, **, P < 0.01, ***, P < 0.001, ****, P < 0.0001, n. s., not significant.

Consistently, the S and N proteins have lower correlations with the non-structural/accessory proteins, while the non-structural/accessory proteins were clustered together. In addition, several sub-clusters were shown among the non-structural/accessory proteins. Interestingly, NSP8 and RdRp have a high correlation (**Figure 2d**). It is known that RdRp, NSP8 and NSP7 could form a tight complex^21,22^, which might contribute to the high correlation. However, the correlation between NSP8 and NSP7 is less significant (**Figure 2c, e**). The structure of the complex shows NSP7 physically connect to RdRp and NSP8, but with most of the protein surface blocked (**Figure 2f**), while NSP8 and RdRp are more accessible. However, NSP7 elicit antibody in a higher frequency (**Figure 1a**), suggesting NSP7 might mainly exist in other forms rather than complex with NSP8 and RdRp, thus has other yet to be discovered biological function(s). In addition, the IgG responses of NSP2 and NSP16 also have a high correlation (**Figure 2c, d-g**). It was reported that PPI (protein-protein interaction) was detected between NSP2 and NSP16^23^. However, we did not detect any direct binding signals between NSP2 and NSP16 *in vitro* (data not shown), suggesting NSP2 and NSP16 might form a complex through the bridging of other proteins. In addition, differences of the response frequencies among the proteins are not associated with the *in vivo* protein expression level ^24^ and the protein length (**Figure S2**).

### IgG responses are associated with severity

It is known that IgG responses against S and N proteins are associated with disease severity and clinical features ^12,14^, however, the correlations to other SARS-CoV-2 proteins, especially non-structural and accessory proteins haven’t been revealed yet. As described above, we divided the patient population into three groups, *i*.*e*. Non-severe, severe survivors and severe non-survivors. Two statistical methods were applied to assess the correlations. One is to analyze the positive rate of IgG against each protein, and the other is to compare the signal intensity distribution among groups (**Figure 3**). For both S1 and N, the overall signals for severe groups were slightly higher than that of non-severe group, but there was no significant difference between the survivors and non-survivors. In contrast, the non-structural/accessory proteins, that with high positive rates/ signal intensities, are more significantly correlated with severity. It is worth noting that for the 6 non-structural and accessory proteins, both the positive rates and signal intensities are significantly higher as the disease exacerbates to more severe stages (**Figure 3)**. These results indicate that the IgG responses against non-structural/accessory proteins are of higher correlations with the disease severity, and may could serve as a better predictor of COVID-19 severity than that of S1 and N proteins.

**Figure 3.**
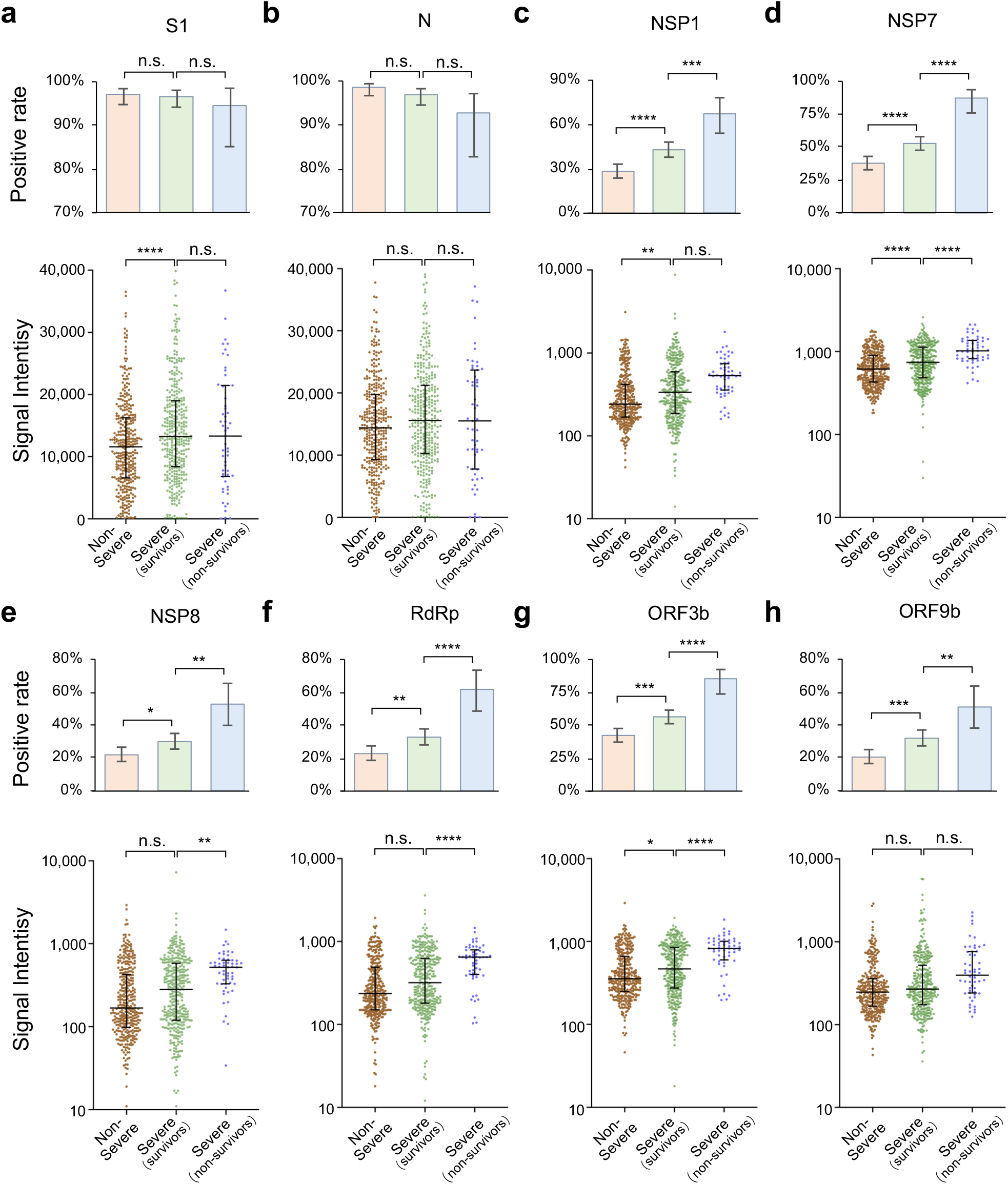
IgG responses are associated with disease severity. **a-h**. IgG positive rate and signal intensity distribution among three patient groups, *i*.*e*., non-severe, severe (survivors) and severe (non-survivors) patients for S1 (**a**), N protein (**b**), NSP1 (**c**), NSP7 (**d**), NSP8 (**e**), RdRp (**f**) ORF3b (**g**) and ORF9b (**h**). For positive rate analysis, error bar was given as the 95% confidential interval. *P*-value was calculated by two-sided χ^2^ test. *, P < 0.05, **, P < 0.01, ***, P < 0.001, ****, P < 0.0001, n. s., not significant. For signal intensity analysis, the middle line was median value; the upper and lower hinges were the values of 75% and 25% percentile. *P-*value was calculated by two-sided *t* test.

We next analyzed the correlation between antibody response and clinical parameter. The clinical parameters, which have statistical correlations with IgG against S1, N, NSP7, NSP8, RdRp, ORF3b and ORF9b, were determined (**Table S3, Figure S3**). All of these parameters are related with severity, suggesting severity is a major factor and confounder that contribute to the correlations. Interestingly, thrombocytopenia, is related with clinical outcome but not severity (**Table 1**), is significantly correlated with NSP7 and ORF3b but not S1 and N, further confirming the higher correlations among antibodies of non-structural/accessory proteins and clinical outcome.

### IgG responses of S1 and N proteins decrease several days before death in non-survivors

The preserve of high tiers of neutralizing antibodies is essential for protecting the patients from re-infection and vaccine development. One critical question is that how long the antibodies against SARS-CoV-2 can last. A recent study found that antibody titers did not decline within 4 months after diagnosis ^25^, while another studies observed rapid decay of antibodies in mild patients ^26,27^. So we analyzed the antibody dynamics with our sample set in which the sera were collected from 0 to about 60 days after initial symptom onset. The seroprevalence or positive rates for both S1 and N reached plateau at about 20 days after symptom onset and maintained afterwards for at least two months for all the three groups (**Figure 4a, b**). However, with regard to signal intensity, a dramatic decrease was observed for the non-survivor group, while not for other two groups, though a slight decline was observed for severe-survivor group (**Figure 4c, d**). The sharp decline in non-survivors might be related to death. To confirm this possibility, we analyzed 35 patients with serum available 0-2 days before death, and 108 survivors with serum available 0-2 day before discharge as control. For each patient, we defined the relative signal for each sample to the sample immediately before death or discharge. Overall, the relative signals declined gradually during the disease progression from about 10 days before death (**Figure 4e**) for non-survivors though the trend differed among individuals. In contrast, there is no significant change for the survivors (**Figure 4f**). These observations might imply a collapse in SARS-CoV-2 related humoral immune in a majority of patients before death and further study are needed to confirm this.

**Figure 4.**
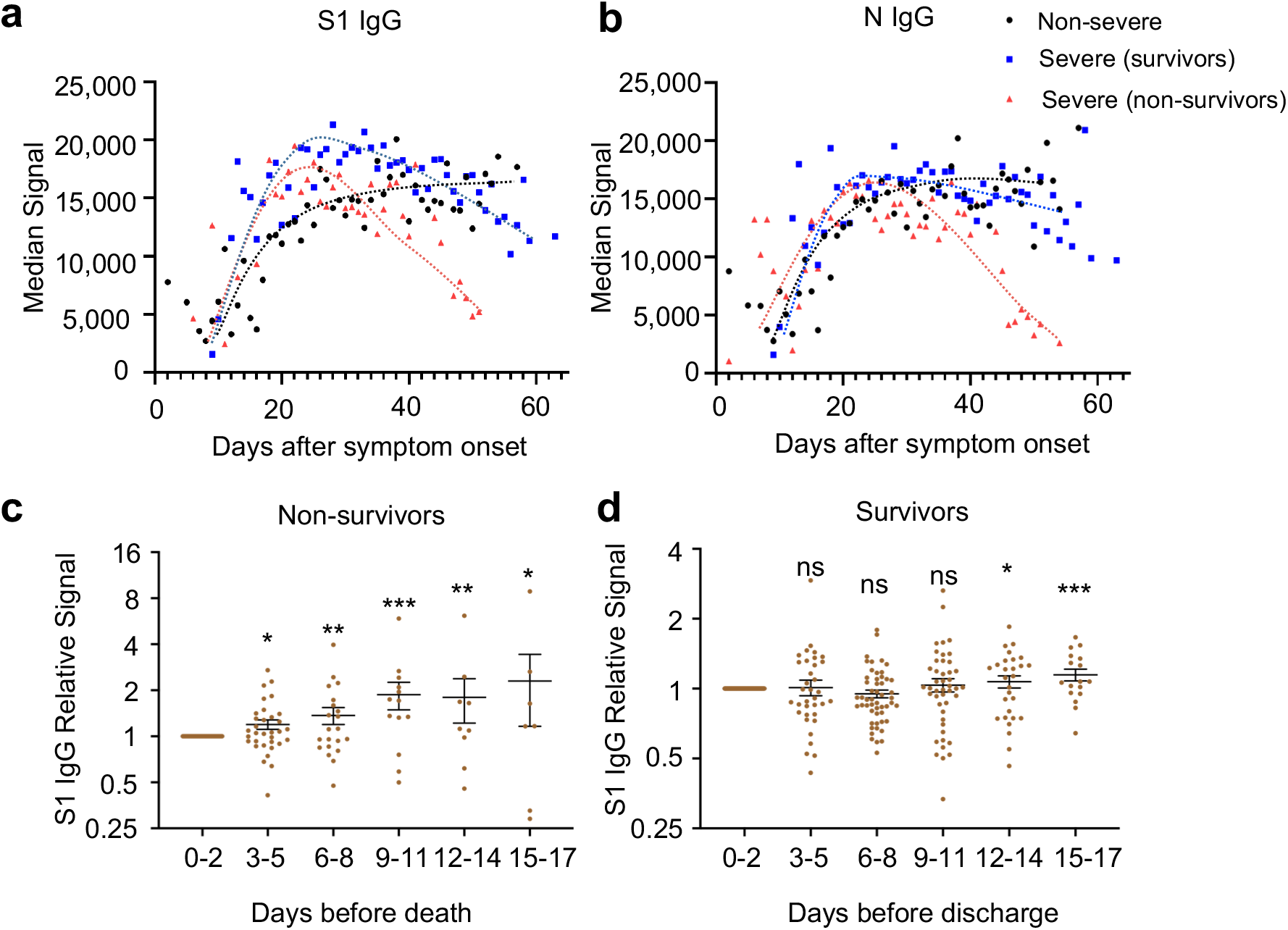
S1 and N IgG decrease several days before death in non-survivors. **a-b**. The trends of median signal intensities of IgG at different time points for S1 (**a**) and N (**b**), among three sample groups, *i. e*., non-severe, severe (survivors) and severe (non-survivors). Samples were grouped per day and the time points with sample number less than 4 were excluded due to lack of statistical significance. **c-d**. Relative S1-IgG signal levels were calculated for each patient, by dividing the signal intensity of the samples collected at other time points vs. samples collected at 0-2 days before the death of non-survivors (**c**, n = 35) or the discharge of survivors (**d**, n = 108). The samples were grouped per three days. For each patient, the signals were averaged if there were more than one sample during each three-day. *P-*value was calculated by two-sided *t* test between the indicated group and the first group (0 - 2 days). *, P < 0.05, **, P < 0.01, ***, P < 0.001, n. s., not significant.

### IgG against the 6 non-structural/accessory proteins decline rapidly during COVID-19 progression

We next analyzed the dynamic of IgG responses for the 6 non-structural/accessory proteins. Surprisingly, the IgG responses, *i*.*e*., the signal intensities and positive rates, against the 6 proteins reached plateau in all the three groups at about 20 days after the symptom onset, and then decreased rapidly for all the three groups (**Figure 5a, 5b**), this is largely different from S1 and N protein (**Figure 4**). We next selected NSP7 IgG as an example for further analysis and depicted the change for each patient (**Figure 5c-e**). Continuous and dramatic decline of IgG against NSP7 for most patients were observed (**Figure 5d, 5e**). These results imply the B cells that producing IgG antibodies against non-structural/accessory proteins might be short-lived, and/ or the underlying mechanism of generating IgG antibodies against non-structural/accessory proteins may differ from that of S1 and N proteins.

**Figure 5.**
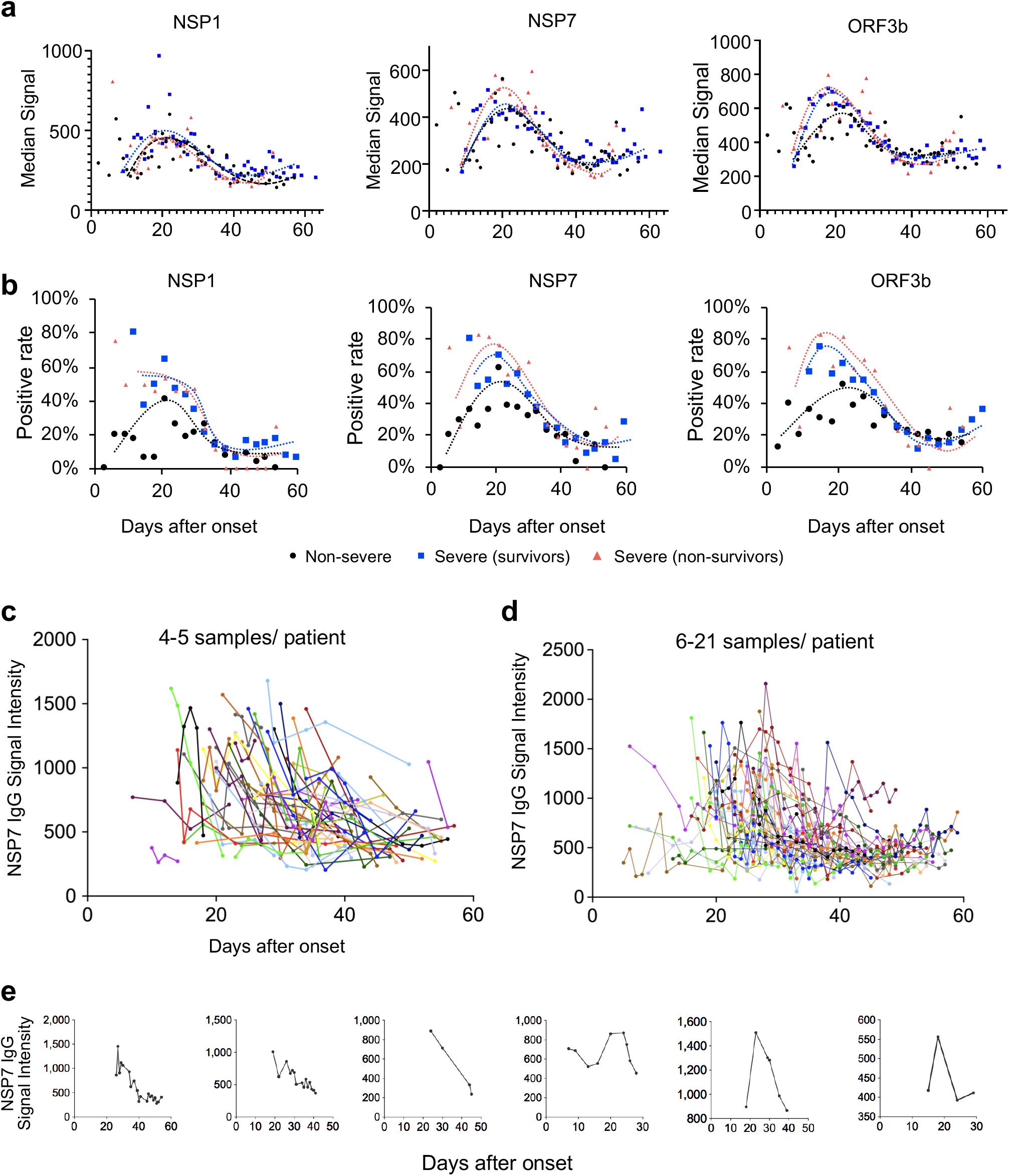
Antibody responses against non-structural proteins and accessory proteins decrease rapidly after 20 days of symptom onset. **a**. The trends of median signal intensities of IgG at different time points for NSP1, NSP7 and ORF3b, among three samples groups, *i. e*., non-severe, severe (survivors) and severe (non-survivors). Samples were grouped per day and the points with sample number less than 4 were excluded. **b**. The trends of positive rate of IgG at different time points for NSP1, NSP7 and ORF3b, among three samples groups, *i. e*., non-severe, severe (survivors) and severe (non-survivors). Samples were grouped per three days. **c-e**. NSP7-IgG signal dynamic changes for the patients with 4-5 samples (**c**) or more samples (**d**) or for some representative individuals (**e**). Each line represents one person.

## Discussion

In this study, we profiled 2,360 sera from 783 COVID-19 patients and 601 control sera using a SARS-CoV-2 proteome microarray. We found that 6 non-structural/accessory proteins elicit strong antibody responses in COVID-19 patients, including NSP1, NSP7, NSP8, RdRp, ORF3b and ORF9b.

It is broadly reported that a batch of clinical laboratory parameters are associated with disease severity of COVID-19 patients, such as lymphopenia, neutrophilia, increased value of CRP, LDH and D-dimer^18–20^. What we observed are mostly consistent with these reports. We also found some parameters that were more related with clinical outcome. For example, there is no obvious difference of the levels of thrombocytopenia between non-severe and severe survivors, but significant decline in non-survivors, suggesting the issue of blood coagulation in COVID-19 patients should be carefully monitored during therapy and fully evaluation for the contribution to death.

NSP1 is a major virulence factor that binds with the small subunit of host ribosome to suppress host gene expression^28^. NSP7, NSP8 and RdRp (NSP12) form a complex, which is involved in the replication and transcription of the SARS-CoV-2 genome, playing an essential role for virus replication. RdRp is the target of a promising drug Remedesivir^22^. ORF3b is reported to be a potent IFN antagonist^29^ and was identified to elicit antibodies in COVID-19 patients^30^. ORF9b can suppress type I IFN production by targeting host protein TOM70 ^31^. Previous studies have found the presence of antibodies against SARS ORF9b^32,33^, and ORF9b IgG antibody identified in convalescent sera, though from a small cohort of sera^14^. However, to our knowledge, all of these findings are based on small cohorts of samples. Here, by analyzing a large cohort of samples, we actually constructed an antibody response landscape of SARS-CoV-2 proteome. This landscape extent our knowledge of the interaction between SARS-CoV-2 and the immune system. However, due to the difficulty of protein preparation, there are still some proteins that are missing on the SARS-CoV-2 proteome microarray, such as ORF8, which has recently been reported to be able to elicit strong antibody response^30^. Some of these missing proteins will be added when we update the microarray.

Comparison of the IgG responses among the antigenic proteins revealed the possibility that the generation of antibodies against the non-structural/accessory proteins are not independent with each other, which means for one patient that positive for one protein tend to has a significant chance to be positive for other proteins. This is expected since these proteins may be simultaneously exposed to or not to the immune system. Interestingly, high correlations were shown between some particular proteins, likely revealing the high associations of these proteins during infection and disease progression. For instance, it is known that RdRp, NSP7 and NSP8 form a complex for replication and translation of the viral genome. Correspondingly, high correlations of their elicited antibody responses, particular for NSP8 and RdRp, were observed. We also observed high correlations between NSP2 and NSP16, suggesting the two proteins might associate *in vivo*.

The function of the antibodies to non-structural/accessory proteins are still largely unknown, our results reveal that the antibody levels are more associated with disease severity, particularly with the final outcome. These findings imply that the antibodies against non-structural/accessory proteins may play more important roles, thus worth further and in-depth investigation. One concern about the antibody against S protein is the possible ADE (antibody dependent enhancement)^34^ which causes uncontrolled release of proinflammatory cytokines, such as IL-1, IL-6, IFN-γ, and TNF-α^35,36^. Meanwhile, the severity of COVID-19 is highly associated with cytokine release syndrome or cytokine storm^36,37^. One possible role of these antibodies against the non-structural/accessory proteins might be to trigger production of more cytokines when they bind the released antigens from the infected cells.

It is still controversial about the duration of the protective antibodies in patients^25,26^. Our data reveal that there is no significant decline of IgG antibodies against S or N protein for mild and severe survivors within 60 days after symptom onset. While the IgG antibodies against non-structural/accessory proteins rapidly decline when they reach the plateau about 20 days after symptom onset. This may could be explained by the gradual decline of virus load which is usually detectable around about 20 days after symptom onset^38,39^. A recently published study identified ORF3b can elicit antibodies and the antibodies can last for two to three months^30^. In fact, for most of the patients, an obvious trend of decline is observed. The duration of antibody is largely dependent on the type of the corresponding B cells or antibody secreting plasma cells, either long-lived or short lived^40,41^. The obvious short life time of the antibodies against non-structural/accessory proteins might due to the suppressed production of long-lived B cells or tend to generate short-lived B cells with unknown mechanism^40^. It seems that there are two distinct mechanisms through which the proteins of SARS-CoV-2 elicit host humoral immune responses: 1) Viral particle is involved as antigen resource, specifically S and N proteins, which elicit potent antibody responses and tend to generate long-lived B cells. These antibodies mainly play a protective role. 2) The infected cell is involved as antigen resource with non-structural/accessory proteins, which elicit weaker antibody responses and tend to be suppressed to generate long-lived B cells. These antibodies might be stronger to induce cytokines to contribute severer outcomes. However, this hypothesis should be confirmed by further studies.

In contrast to the antibody levels of S and N protein are stable for survivors, we observed the overall antibody levels start to decline in non-survivors at about ten days before death. This observation implies the possible protective function of these antibodies in patients, and a collapse of humoral immune might occur for most patients immediately before death. Further studies are needed to confirm this observation with more samples.

Taken together, we revealed a comprehensive antibody landscape against SARS-CoV-2 proteome. The results were assured by a large cohort of 2,360 COVID-19 sera. Distinct characteristics of the antibodies against non-structural/accessory proteins and structural proteins were shown for the first time, with regard to patterns of antibody responses, associations with severity/outcome, and the dynamic. We strongly believe that the antibody landscape revealed in this study will facilitate a deeper understanding the immunity of SARS-CoV-2, predict the final outcome, may provide potential biomarkers for precise monitoring of COVID-19 progression, and may guide the development of effective vaccines.

## Data Availability

The SARS-CoV-2 proteome microarray data are deposited on Protein Microarray Database under the accession number PMDE243 (http://www.proteinmicroarray.cn). Additional data related to this paper may be requested from the authors.

## Acknowledgments

We thank Dr. Daniel M. Czajkowsky for English editing and critical comments. We thank Prof. H. Eric Xu (Shanghai Institute of Materia Medica) for providing RdRp protein and Dawei Shi from National Institutes for Food and Drug Control, Beijing, China for providing negative reference control samples. We also thank Healthcode Co., Ltd., Hangzhou Bioeast biotech Co., Ltd. and VACURE Biotechnology Co., Ltd. for providing reagents. This work was partially supported by the National Key Research and Development Program of China Grant (No. 2016YFA0500600), National Natural Science Foundation of China (No. 31970130, 31600672, 31670831, 31370813, 31900112 and 21907065).

## Author contributions

S-C. T. and X-L. F. developed the conceptual ideas and designed the study. Y.L., Z-W.X., Q.L., D-Y.L., Z-W.X., M-L. M., B.Z., H.C., C-Z.Y., J-B.X., X-N.W., Y-X.Z., H-N.Z., H-W.J., H.Q. Y-D.Z., X-S.L., Z-J.Y. and S-J.G. performed the experiments and data analysis.. Z-Y.S., F.W., H-Y.H., Y-D.Z., X-S. L., Z-J.Y., H-M.S., J-X.W., L-Y.C., S-Q.L., P-F.P., and H.S. collected the sera samples. S-C.T. and Y.L. wrote the manuscript with suggestions from other authors.

## Declaration of interests

The authors declare no competing interests.

## Methods and Materials

### Protein microarray fabrication

The SARS-CoV-2 proteome microarray used this study is a updated version of the original one^14^. Three more proteins, *i. e*., ORF3a, ORF3b and ORF7b, were expressed by ourself and added. The 4^th^ protein, RdRp was provided by H. Eric Xu^22^. The protein microarray was fabricated as previously described^14^. Briefly, the proteins with indicated concentrations, along with the negative (GST, Biotin-control and eGFP) and positive controls (Human IgG, Human IgM and ACE2-Fc), were printed in quadruplicate on PATH substrate slide (Grace Bio-Labs, Oregon, USA) to generate identical arrays in a 2× 7 subarray format using Super Marathon printer (Arrayjet, UK). The microarrays were used for serum profiling as described previously with minor modifications^19^. Protein microarrays were stored at −80°C until use.

### Patients and samples

The study was approved by the Ethical Committee of Tongji Hospital, Tongji Medical College, Huazhong University of Science and Technology, Wuhan, China (ITJ-C20200128). Written informed consent was obtained from all participants enrolled in this study. All COVID-19 patients were laboratory confirmed and hospitalized during the period from 25 January 2020 and 28 April 2020. Sera of the control group from healthy donors, lung cancer patients, patients with autoimmune diseases were collected from Ruijin Hospital, Shanghai, China or Tongren Hospital, Shanghai, China. The negative reference samples were from National Institutes for Food and Drug Control. All the samples were stored at −80°C until use.

### Microarray-based serum analysis

A 14-chamber rubber gasket was mounted onto each slide to create individual chambers for the 14 identical subarrays. The microarray was used for serum profiling as described by Li, *et al*.^39^ with minor modifications. Briefly, the arrays stored at −80°C were warmed to room temperature and then incubated in blocking buffer (3% BSA in 1×PBS buffer with 0.1% Tween 20) for 3 h. Serum samples were diluted 1: 200 in PBS containing 0.1% Tween 20, added with 0.5 mg/mL^-1^ total *E. coli* lysate. A total of 200 μL of diluted serum or buffer only was incubated with each subarray overnight at 4°C. The arrays were washed with 1 × PBST and the signals were readout by incubating with Cy3-conjugated goat anti-human IgG and Alexa Fluor 647-conjugated donkey anti-human IgM (Jackson ImmunoResearch, PA, USA). These two fluorescent conjugated antibodies were diluted 1: 1,000 in 1 × PBST and incubated at room temperature for 1 h. The microarrays were then washed with 1 × PBST, dried by centrifugation at room temperature, scanned by LuxScan 10K-A (CapitalBio Corporation, Beijing, China) with the parameters set as 95% laser power/ PMT 550 and 95% laser power/ PMT 480 for IgM and IgG, respectively. The fluorescent intensity data was extracted by GenePix Pro 6.0 software (Molecular Devices, CA, USA). A pool of 50 randomly selected patient sera was used as a standard positive control. Block #14 of each slide was incubated with the positive control. Data normalization among slides was performed by a linear method with the data from the positive control. Specifically, a normalization factor for each slide was calculated by a liner regression function of the signals of the positive control of the given slide with the averaged signals of all slides, and then the signals of all the proteins from the slide were divided by the factor.

### Statistics

Signal Intensity was defined as the median of the foreground subtracted by the median of background for each spot and then the quadruplicate spots were averaged for each protein. IgG and IgM data were analyzed separately. Pearson correlation coefficient between two proteins or indicators and the corresponding *p*-value was calculated by SPSS software under the default parameters. Cluster analysis was performed by pheatmap package of R ^42^. To calculate the positive rate of antibody response for each protein, mean + 2 x standard deviation (SD) of the control sera were used to set the threshold.

## Supplemental Information

**Figure S1.**
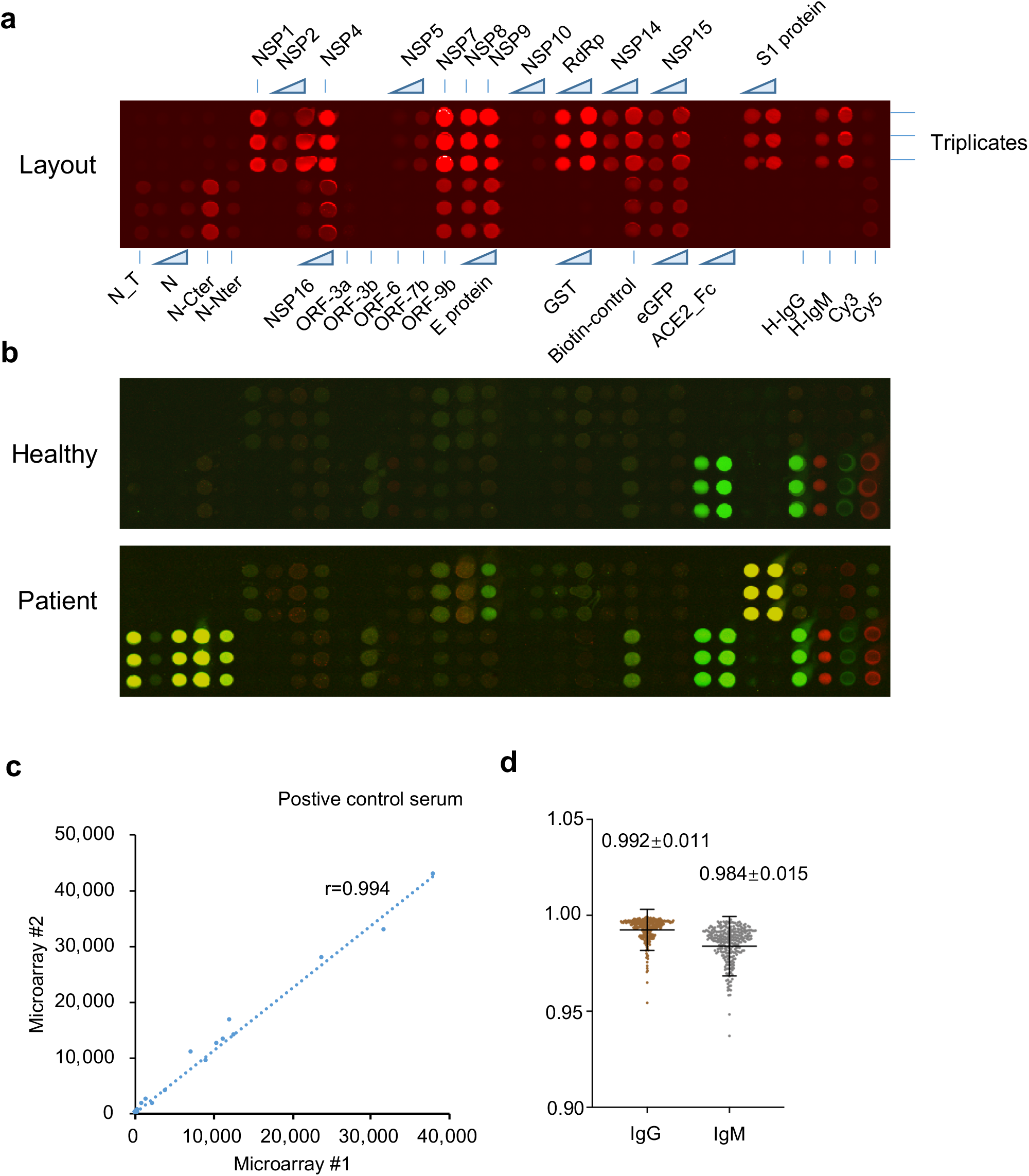
SARS-CoV-2 proteome microarray and the assessment of (reproducibility (related to Figure 1). **a**. The layout of the SARS-CoV-2 proteome microarray. The locations of proteins and controls are indicated. **b**. Representative images of the microarray screened by sera from a healthy control and a COVID-19 patient. **c**. Correlation analysis between two microarrays probed independently with a positive control serum. **d**. Statistical analysis of the Pearson correlation coefficients between the microarrays incubated with the positive control serum with the averaged data set (see methods). The data are present as mean + SD.

**Figure S2.**
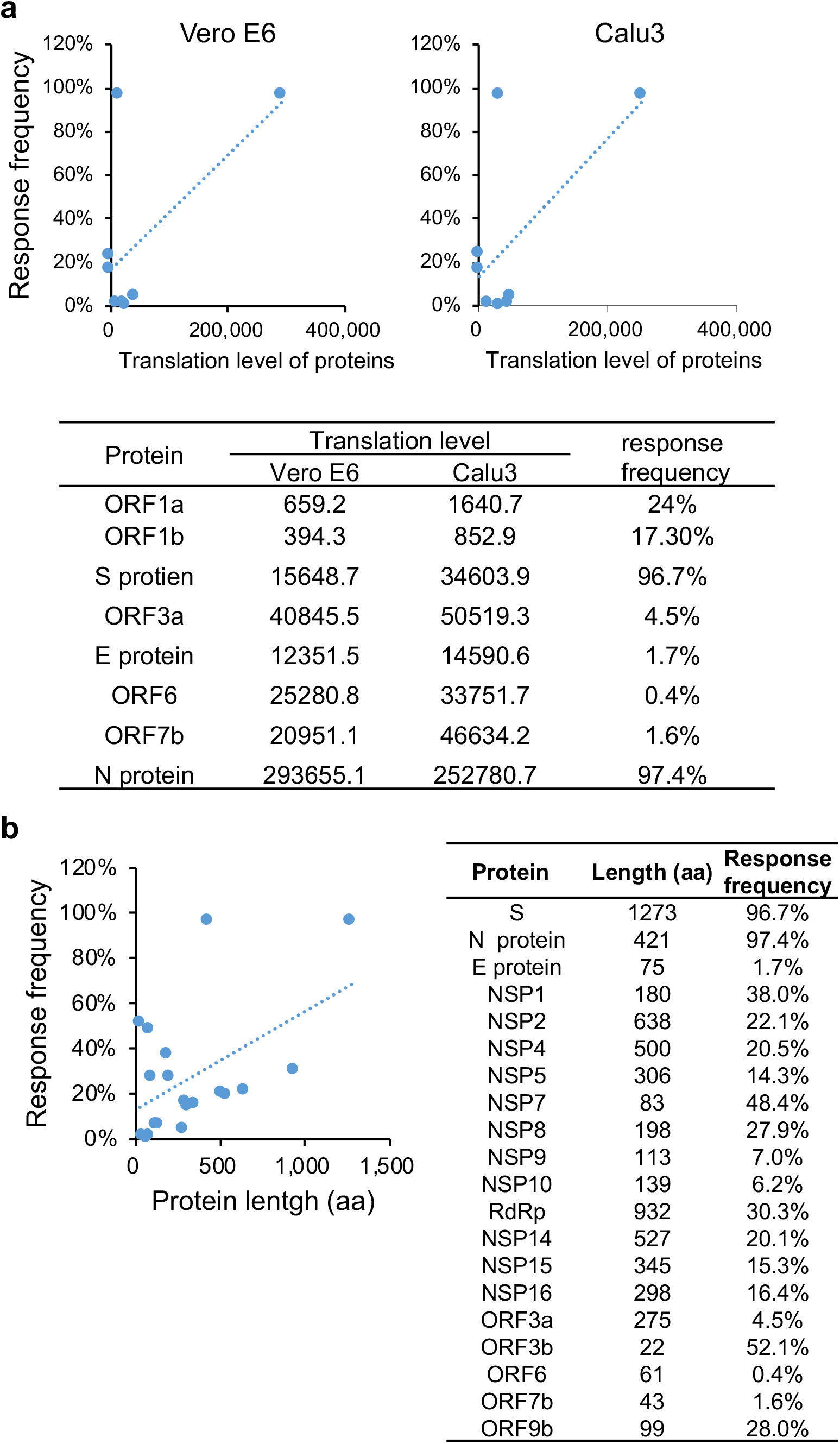
Antibody responses are not associated with protein abundance or length (related to figure 2). **a-b**. Correlations between antibody response frequency and protein abundance (Finkel et al., 2020) (**a**), and protein length (**b**).

**Figure S3.**
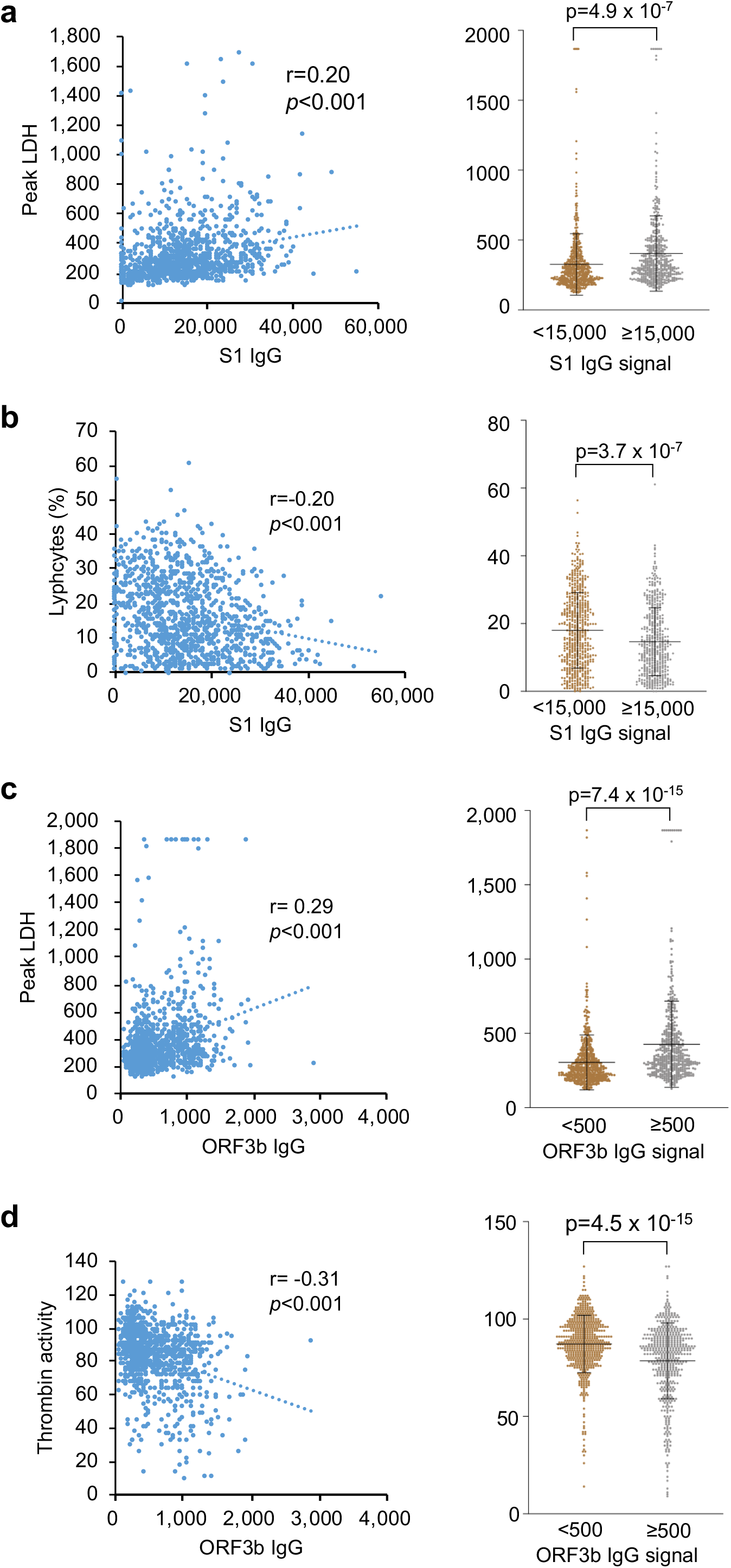
IgG responses are associated with clinical parameters (related to figure 3). **a-d**. Correlations and statistical analysis of IgG response against indicate proteins and clinical parameters. The right part for each panel depicts the distribution of the values for corresponding clinical parameter in lower and higher IgG response groups. *P*-values were calculated with two-sided *t* test.

**Table S1.** Serum Samples and patients (related to Figure 1)

| Group |  | COVID-19 | Control-1 | Control-2 |
| --- | --- | --- | --- | --- |
| Patients (n) |  | 783 | 469 | 73 |
| Serum samples (n) |  | 2,360 | 469 | 73 |
| Patients with samples >14 days after onset |  | 756 | - | - |
| Age |  | 61.4±14.5 | 53.7±20.6 | N/A |
| Gender | Male | 377 | 224 | N/A |
|  | Female | 379 | 245 | N/A |
| Severity/ outcome | non-severe | 347 |  |  |
|  | Severe (survivors) | 354 | - | - |
|  | Severe (non-survivors) | 55 |  |  |
| Source |  | Tongji Hospital, Wuhan | Tongren Hospital, Shanghai<br>Ruijin Hospital, Shanghai | National Institutes for Food and Drug Control, Beijing |
| Subtype and number |  | - | Healthy: 92;<br>URI: 104; AID: 120;<br>LC: 41;<br>Other diseases: 112 | Negative reference samples |

**Table S2.**
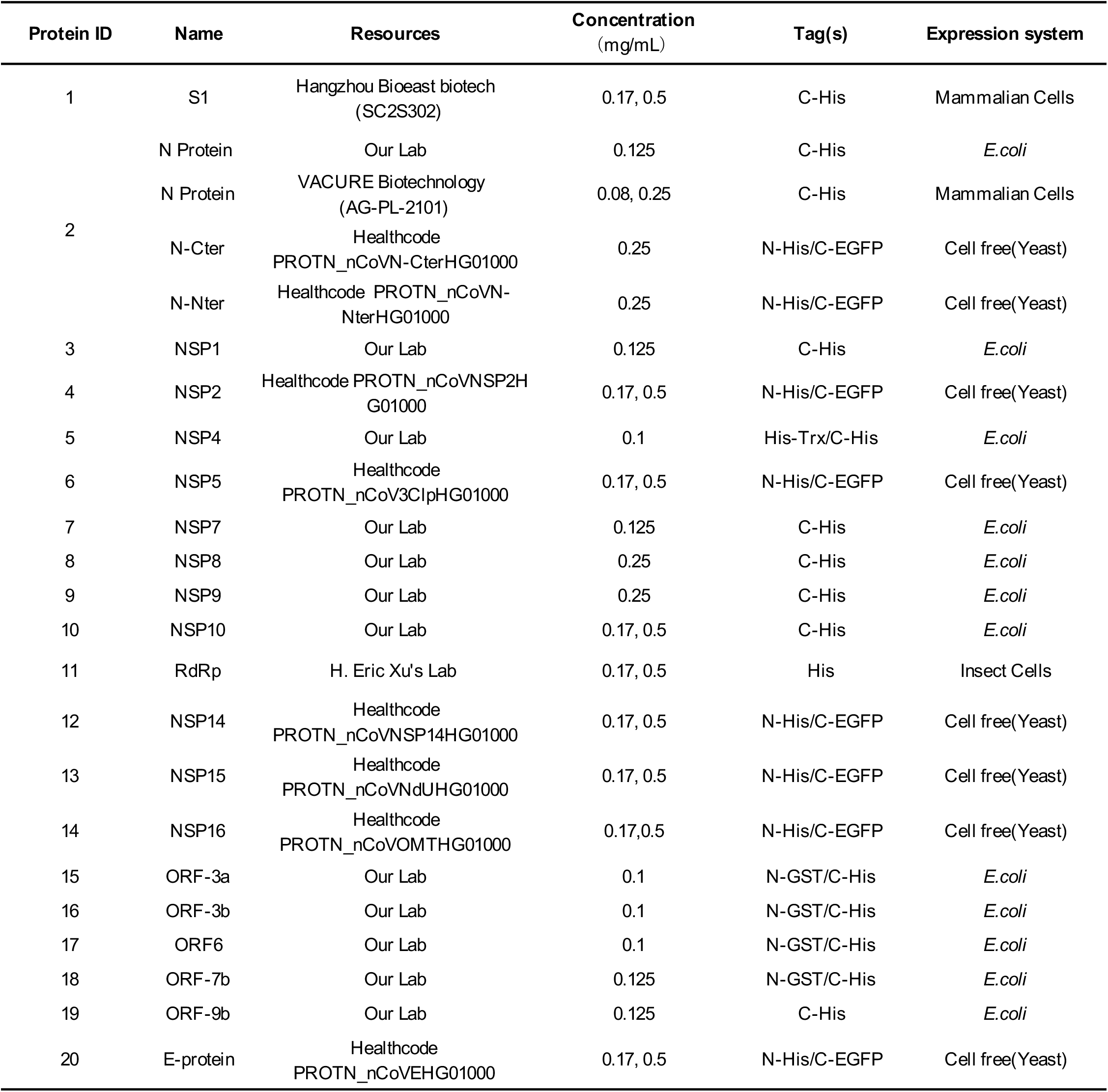
SARS-CoV-2 proteins included in the proteome microarray (related to Figure 1 and Figure S1)

**Table S3.**
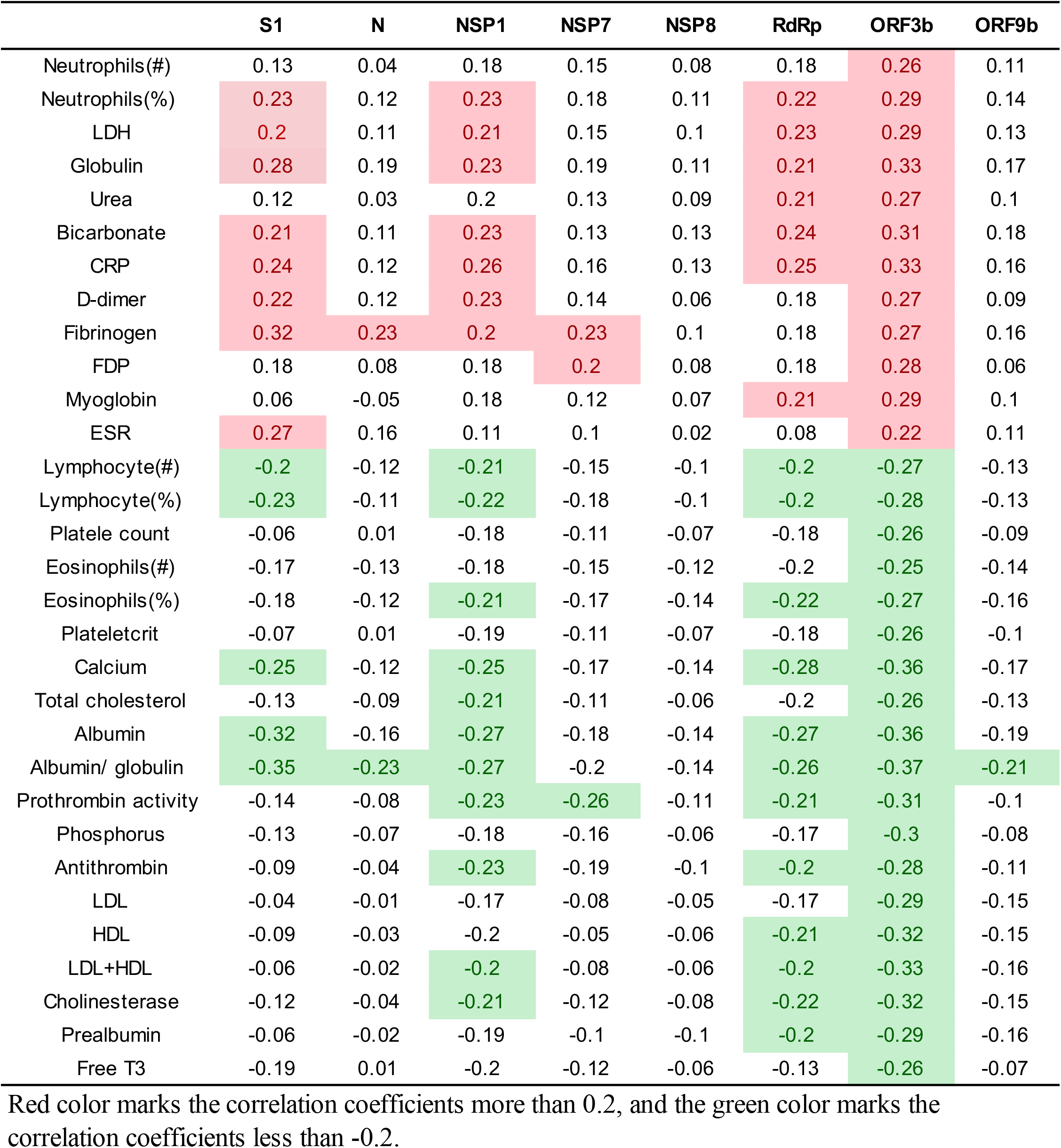
IgG responses are associated with clinical parameters (related to Figure 3 and Figure S3)

|  | S1 | N | NSP1 | NSP7 | NSP8 | RdRp | ORF3b | ORF9b |
| --- | --- | --- | --- | --- | --- | --- | --- | --- |
| Neutrophils(#) | 0.13 | 0.04 | 0.18 | 0.15 | 0.08 | 0.18 | 0.26 | 0.11 |
| Neutrophils(%) | 0.23 | 0.12 | 0.23 | 0.18 | 0.11 | 0.22 | 0.29 | 0.14 |
| LDH | 0.2 | 0.11 | 0.21 | 0.15 | 0.1 | 0.23 | 0.29 | 0.13 |
| Globulin | 0.28 | 0.19 | 0.23 | 0.19 | 0.11 | 0.21 | 0.33 | 0.17 |
| Urea | 0.12 | 0.03 | 0.2 | 0.13 | 0.09 | 0.21 | 0.27 | 0.1 |
| Bicarbonate | 0.21 | 0.11 | 0.23 | 0.13 | 0.13 | 0.24 | 0.31 | 0.18 |
| CRP | 0.24 | 0.12 | 0.26 | 0.16 | 0.13 | 0.25 | 0.33 | 0.16 |
| D-dimer | 0.22 | 0.12 | 0.23 | 0.14 | 0.06 | 0.18 | 0.27 | 0.09 |
| Fibrinogen | 0.32 | 0.23 | 0.2 | 0.23 | 0.1 | 0.18 | 0.27 | 0.16 |
| FDP | 0.18 | 0.08 | 0.18 | 0.2 | 0.08 | 0.18 | 0.28 | 0.06 |
| Myoglobin | 0.06 | -0.05 | 0.18 | 0.12 | 0.07 | 0.21 | 0.29 | 0.1 |
| ESR | 0.27 | 0.16 | 0.11 | 0.1 | 0.02 | 0.08 | 0.22 | 0.11 |
| Lymphocyte(#) | -0.2 | -0.12 | -0.21 | -0.15 | -0.1 | -0.2 | -0.27 | -0.13 |
| Lymphocyte(%) | -0.23 | -0.11 | -0.22 | -0.18 | -0.1 | -0.2 | -0.28 | -0.13 |
| Platele count | -0.06 | 0.01 | -0.18 | -0.11 | -0.07 | -0.18 | -0.26 | -0.09 |
| Eosinophils(#) | -0.17 | -0.13 | -0.18 | -0.15 | -0.12 | -0.2 | -0.25 | -0.14 |
| Eosinophils(%) | -0.18 | -0.12 | -0.21 | -0.17 | -0.14 | -0.22 | -0.27 | -0.16 |
| Plateletcrit | -0.07 | 0.01 | -0.19 | -0.11 | -0.07 | -0.18 | -0.26 | -0.1 |
| Calcium | -0.25 | -0.12 | -0.25 | -0.17 | -0.14 | -0.28 | -0.36 | -0.17 |
| Total cholesterol | -0.13 | -0.09 | -0.21 | -0.11 | -0.06 | -0.2 | -0.26 | -0.13 |
| Albumin | -0.32 | -0.16 | -0.27 | -0.18 | -0.14 | -0.27 | -0.36 | -0.19 |
| Albumin/ globulin | -0.35 | -0.23 | -0.27 | -0.2 | -0.14 | -0.26 | -0.37 | -0.21 |
| Prothrombin activity | -0.14 | -0.08 | -0.23 | -0.26 | -0.11 | -0.21 | -0.31 | -0.1 |
| Phosphorus | -0.13 | -0.07 | -0.18 | -0.16 | -0.06 | -0.17 | -0.3 | -0.08 |
| Antithrombin | -0.09 | -0.04 | -0.23 | -0.19 | -0.1 | -0.2 | -0.28 | -0.11 |
| LDL | -0.04 | -0.01 | -0.17 | -0.08 | -0.05 | -0.17 | -0.29 | -0.15 |
| HDL | -0.09 | -0.03 | -0.2 | -0.05 | -0.06 | -0.21 | -0.32 | -0.15 |
| LDL+HDL | -0.06 | -0.02 | -0.2 | -0.08 | -0.06 | -0.2 | -0.33 | -0.16 |
| Cholinesterase | -0.12 | -0.04 | -0.21 | -0.12 | -0.08 | -0.22 | -0.32 | -0.15 |
| Prealbumin | -0.06 | -0.02 | -0.19 | -0.1 | -0.1 | -0.2 | -0.29 | -0.16 |
| Free T3 | -0.19 | 0.01 | -0.2 | -0.12 | -0.06 | -0.13 | -0.26 | -0.07 |

Red color marks the correlation coefficients more than 0.2, and the green color marks the correlation coefficients less than -0.2.

## Notes

### Competing Interest Statement

The authors have declared no competing interest.

### Clinical Trial

NA

### Author Declarations

The study was approved by the Ethical Committee of Tongji Hospital, Tongji Medical College, Huazhong University of Science and Technology, Wuhan, China (ITJ-C20200128).

## References

1. Zhou, P. et al. A pneumonia outbreak associated with a new coronavirus of probable bat origin. Nature 579, 270–273 (2020).

2. Wu, F. et al. A new coronavirus associated with human respiratory disease in China. Nature 579, 265–269 (2020).

3. Dong, E., Du, H. & Gardner, L. An interactive web-based dashboard to track COVID-19 in real time. Lancet Infect. Dis. 20, 533–534 (2020).

4. Wu, A. et al. Genome Composition and Divergence of the Novel Coronavirus (2019-nCoV) Originating in China. Cell Host Microbe 27, 325–328 (2020).

5. Ge, X.-Y. et al. Isolation and characterization of a bat SARS-like coronavirus that uses the ACE2 receptor. Nature 503, 535–538 (2013).

6. Wrapp, D. et al. Cryo-EM structure of the 2019-nCoV spike in the prefusion conformation. Science (80-.). 367, 1260–1263 (2020).

7. Yan, R. et al. Structural basis for the recognition of SARS-CoV-2 by full-length human ACE2. Science (80-.). 367, 1444–1448 (2020).

8. Lan, J. et al. Structure of the SARS-CoV-2 spike receptor-binding domain bound to the ACE2 receptor. Nature 581, 215–220 (2020).

9. Hoffmann, M. et al. SARS-CoV-2 Cell Entry Depends on ACE2 and TMPRSS2 and Is Blocked by a Clinically Proven Protease Inhibitor. Cell 181, 271-280.e8 (2020).

10. Shrock, E. et al. Viral epitope profiling of COVID-19 patients reveals cross-reactivity and correlates of severity. Science (80-.). doi: 10.1126/science.abd4250 (2020) doi:10.1126/science.abd4250.

11. Jiang, S., Hillyer, C. & Du, L. Neutralizing Antibodies against SARS-CoV-2 and Other Human Coronaviruses. Trends Immunol. 41, 355–359 (2020).

12. Long, Q. X. et al. Antibody responses to SARS-CoV-2 in patients with COVID-19. Nat. Med. 26, 845–848 (2020).

13. Vabret, N. et al. Immunology of COVID-19: Current State of the Science. Immunity 52, 910– 941 (2020).

14. Jiang, H. et al. SARS-CoV-2 proteome microarray for global profiling of COVID-19 specific IgG and IgM responses. Nat. Commun. 11, 3581 (2020).

15. Wang, H. et al. SARS-CoV-2 proteome microarray for mapping COVID-19 antibody interactions at amino acid resolution. ACS Cent. Sci. DOI: dx.doi.org/10.1021/acscentsci.0c00742 (2020) doi:10.1101/2020.03.26.994756.

16. Li, Y. et al. Longitudinal serum autoantibody repertoire profiling identifies surgery-associated biomarkers in lung adenocarcinoma. EBioMedicine 53, 102674 (2020).

17. Lei, Q. et al. Antibody dynamics to SARS-CoV-2 in asymptomatic COVID-19 infections. Allergy doi: 10.1111/all.14622 (2020).

18. Wu, C. et al. Risk Factors Associated With Acute Respiratory Distress Syndrome and Death in Patients With Coronavirus Disease 2019 Pneumonia in Wuhan, China. JAMA Intern. Med. 180, 934–943 (2020).

19. García, L. F. Immune Response, Inflammation, and the Clinical Spectrum of COVID-19. Front. Immunol. 11, 1441 (2020).

20. Huang, C. et al. Clinical features of patients infected with 2019 novel coronavirus in Wuhan, China. Lancet 6736, 1–10 (2020).

21. Gao, Y. et al. Structure of the RNA-dependent RNA polymerase from COVID-19 virus. Science (80-.). 368, 779–782 (2020).

22. Yin, W. et al. Structural basis for inhibition of the RNA-dependent RNA polymerase from SARS-CoV-2 by remdesivir. Science (80-.). 368, 1499–1504 (2020).

23. Li, J. et al. Virus-Host Interactome and Proteomic Survey Reveal Potential Virulence Factors Influencing SARS-CoV-2 Pathogenesis. Med DOI: 10.1016/j.medj.2020.07.002 (2020) doi:10.1016/j.medj.2020.07.002.

24. Finkel, Y. et al. The coding capacity of SARS-CoV-2. Nature doi: 10.1038/s41586-020-2739-1 (2020) doi:10.1038/s41586-020-2739-1.

25. Gudbjartsson, D. F. et al. Humoral Immune Response to SARS-CoV-2 in Iceland. N. Engl. J. Med. (2020) doi:10.1056/NEJMoa2026116.

26. Ibarrondo, F. J. et al. Rapid Decay of Anti-SARS-CoV-2 Antibodies in Persons with Mild Covid-19. The New England journal of medicine vol. 383 1085–1087 (2020).

27. Long, Q. X. et al. Clinical and immunological assessment of asymptomatic SARS-CoV-2 infections. Nat. Med. 26, 1200–1204 (2020).

28. Thoms, M. et al. Structural basis for translational shutdown and immune evasion by the Nsp1 protein of SARS-CoV-2. Science (80-.). 369, 1249–1255 (2020).

29. Konno, Y. et al. SARS-CoV-2 ORF3b Is a Potent Interferon Antagonist Whose Activity Is Increased by a Naturally Occurring Elongation Variant. Cell Rep. 32, 108185 (2020).

30. Hachim, A. et al. ORF8 and ORF3b antibodies are accurate serological markers of early and late SARS-CoV-2 infection. Nat. Immunol. 21, 1293–1301 (2020).

31. Jiang, H.-W. et al. SARS-CoV-2 Orf9b suppresses type I interferon responses by targeting TOM70. Cell. Mol. Immunol. 17, 998–1000 (2020).

32. Guo, J.-P., Petric, M., Campbell, W. & McGeer, P. L. SARS corona virus peptides recognized by antibodies in the sera of convalescent cases. Virology 324, 251–256 (2004).

33. Qiu, M. et al. Antibody responses to individual proteins of SARS coronavirus and their neutralization activities. Microbes Infect. 7, 882–889 (2005).

34. Wu, F. et al. Antibody-dependent enhancement (ADE) of SARS-CoV-2 infection in recovered COVID-19 patients: studies based on cellular and structural biology analysis. medRxiv 2020.10.08.20209114 (2020) doi:10.1101/2020.10.08.20209114.

35. Liu, L. et al. Anti-spike IgG causes severe acute lung injury by skewing macrophage responses during acute SARS-CoV infection. JCI insight 4, e123158 (2019).

36. Ragab, D., Salah Eldin, H., Taeimah, M., Khattab, R. & Salem, R. The COVID-19 Cytokine Storm; What We Know So Far. Front. Immunol. 11, 1446 (2020).

37. Mangalmurti, N. & Hunter, C. A. Cytokine Storms: Understanding COVID-19. Immunity 53, 19–25 (2020).

38. Azkur, A. K. et al. Immune response to SARS-CoV-2 and mechanisms of immunopathological changes in COVID-19. Allergy 75, 1564–1581 (2020).

39. He, X. et al. Temporal dynamics in viral shedding and transmissibility of COVID-19. Nat. Med. 26, 672–675 (2020).

40. Nutt, S. L., Hodgkin, P. D., Tarlinton, D. M. & Corcoran, L. M. The generation of antibody-secreting plasma cells. Nat. Rev. Immunol. 15, 160–171 (2015).

41. Cyster, J. G. & Allen, C. D. C. B Cell Responses: Cell Interaction Dynamics and Decisions. Cell 177, 524–540 (2019).

42. Kolde, R. Pheatmap: Pretty Heatmaps. https://cran.r-project.org/web/packages/pheatmap/index.html (2015).

